# Automated and combined HIV, HBV, HCV, and syphilis testing among illegal gold miners in French Guiana using a standardized dried blood device

**DOI:** 10.1101/2022.05.02.22274368

**Authors:** Amandine Pisoni, Elisa Reynaud, Maylis Douine, Louise Hureau, Carmen Alcocer Cordellat, Roxane Schaub, Dennis Poland, Richard Monkel, Joan Lommen, Konstantin Yenkoyan, Stephen Vreden, Mathieu Nacher, Edouard Tuaillon

## Abstract

Blood spotted onto filter paper can be easily collected outside healthcare facilities and shipped to a central laboratory for serological testing. However, dried blood testing generally requires manual processing for pre-analytical steps. In this study, we used a standardized blood collection device combined with an automated elution system to test illegal gold miners living in French Guiana for HIV, HBV, HCV and syphilis. We included 378 participants, 102 females and 266 males, in three illegal gold mining resting sites. Blood collected on the Ser-Col device (Labonovum) was eluted using an automated system (SCAUT Ser-Col automation, Blok System Supply) and an automated analyzer (Alinity i, Abbott). Ser-Col results were compared to both plasma results, considered the gold standard, and to DBS results, considered the reference sampling method using dried blood. In plasma samples, two participants (0.5%) tested positive for HIV, six (1.5%) tested positive for hepatitis B surface antigen (HBsAg), eight were weakly positive for anti-HCV antibodies but negative for HCV RNA, and 47 tested positive for treponemal antibodies (12.4%), including 20 females (19.6%) and 27 males (9.8%, p= 0.010179). We observed a full concordance of Ser-Col and DBS results for HIV diagnosis compared to plasma results. Ser-Col and DBS samples tested positive in five HBsAg carriers and negative for one participant with a low HBsAg level in plasma (0.5 IU/mL). All participants tested negative for HCV in Ser-Col and DBS samples, including the eight participants who tested low positive for HCV antibodies and HCV RNA negative in plasma. Among syphilis seropositive participants, 41 (87.2%) and 40 (85.1%) tested positive for treponemal antibodies in Ser-Col and DBS samples, respectively. The Ser-Col method allows automated dried blood testing of HIV, HBV, HCV and syphilis with performances comparable to DBS. Automated approaches to test capillary blood transported on dried blood devices may facilitate large-scale surveys and improve testing of populations living in remote areas.

**Graphical abstract:** 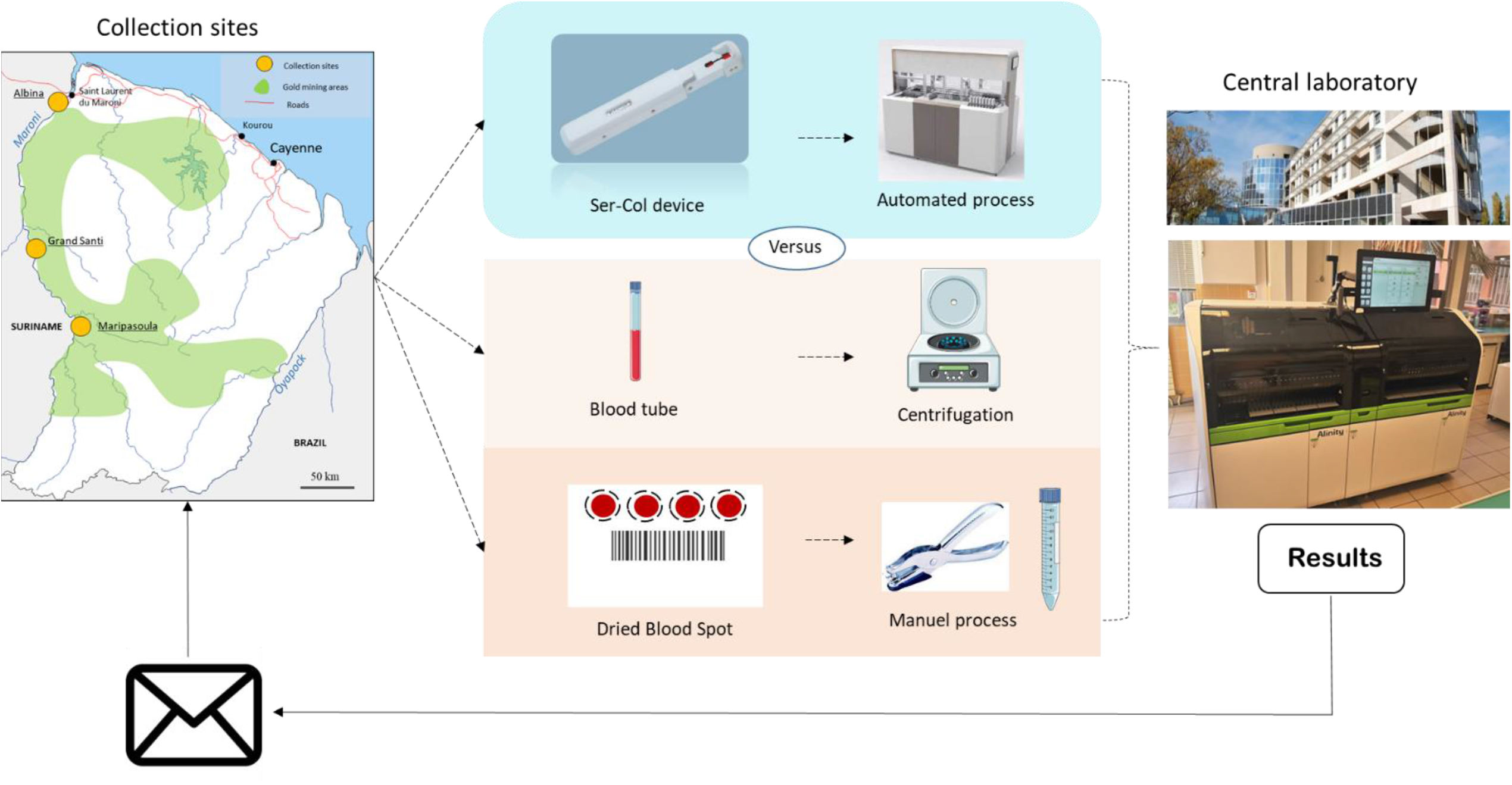

## Introduction

Reducing the burden of infectious diseases requires a global approach that includes underprivileged people. To date, a significant part of the world’s population has insufficient access to reliable infectious disease diagnostic services. Migration, forced displacement of populations, weak infrastructure and long distance from health services are common unfavorable health factors. A lack of in vitro diagnostic tools contributes largely to the mortality and morbidity induced by infectious diseases in resource-limited countries (Organization, 2013). A low number of laboratories and poor logistical infrastructure are major problems in many of these countries. Laboratory equipment defined by WHO as essential for basic service readiness is also insufficient (Leslie et al., 2017). The weakness of human resources - health professionals and biologists - should also be noted (Kruk et al., 2018).

Capillary blood sampling performed outside healthcare facilities is one strategy to improve the screening of hard-to-reach and key populations. Dried blood spot (DBS) testing programs have been implemented for several years, targeting intravenous drug users (McLeod et al., 2014), sex workers (Shokoohi et al., 2016), homeless populations (Foroughi et al., 2017), migrants and men who have sex with men (Bogowicz et al., 2016). Today, WHO considers DBS samples as a possible alternative to serum/plasma to improve access to HIV, HBV and HCV screening and therapeutic monitoring. However, DBS testing has several constraints, such as a lack of standardized blood sampling and elution procedures and the need to manually process DBS cards, which have limited the use of DBS in clinical laboratories.

In French Guiana, about 10,000 illegal gold miners, mainly Brazilians (95%), work on more than 700 different mining sites located in the primary forest (Douine et al., 2017). This neglected population experiences very poor access to Health (Douine et al., 2017). People working on gold panning sites are particularly exposed to vector-borne diseases, including malaria (Douine et al., 2016) and leishmaniasis (Douine et al., 2017). Data also suggest a higher prevalence of sexually transmitted infections such as HIV, hepatitis B (HBV) and syphilis than in other populations of French Guiana (Douine et al., 2019). Transactional sex occurs frequently in gold mining sites (Douine et al., 2019; Gaillet et al., 2017). According to a survey carried out in the city of Cayenne in French Guiana, it may contribute to 45% of HIV cases in men and 10.7% in women (Nacher et al., 2010). In a study carried out on the gold sites of French Guiana, about 10% of women declared having already engaged in sex work activity (Douine et al., 2017).

In this study, we used an innovative blood collection device combined with an automated elution system to test illegal gold miners living in French Guiana for HIV, HBV, HCV and syphilis.

## Material and Methods

### Study design

This study is part of the EU-funded “SCAUT” research project (from finger to laboratory: personalized and automated blood collection for laboratory diagnostics). Participants were adults working in illegal gold mining sites in French Guiana and included in the Malakit study. This study aimed to evaluate an innovative malaria control strategy amongst gold miners working illegally in French Guiana. Participants were included after having provided informed written consent. Testing for HIV, hepatitis B virus (HBV), hepatitis C virus (HCV) and syphilis was offered at three resting sites located along the border between French Guyana and Suriname, on the Maroni river: Albina, Grand Santi and Maripasoula (Fig. 1). All participants had left a mining site less than seven days prior to being tested. Samples collected from 1^st^ October to 16^th^ November were sent from the resting site to the hospital center of Cayenne and then to the Montpellier University Hospital to be tested. Results from plasma samples were communicated to collection sites within two weeks on each site during the post-test counseling. Paired samples of Ser-Col and DBS were tested in parallel for the SCAUT study. The study obtained approval from ethical committee in France: INSERM Ethics Evaluation Committee (No. 14-187 of 09.12.2014), in Suriname: CMWO. Approval No: VG 25-17; ClinicalTrials.gov identifier (NCT number: NCT03695770).

**Figure 1.**
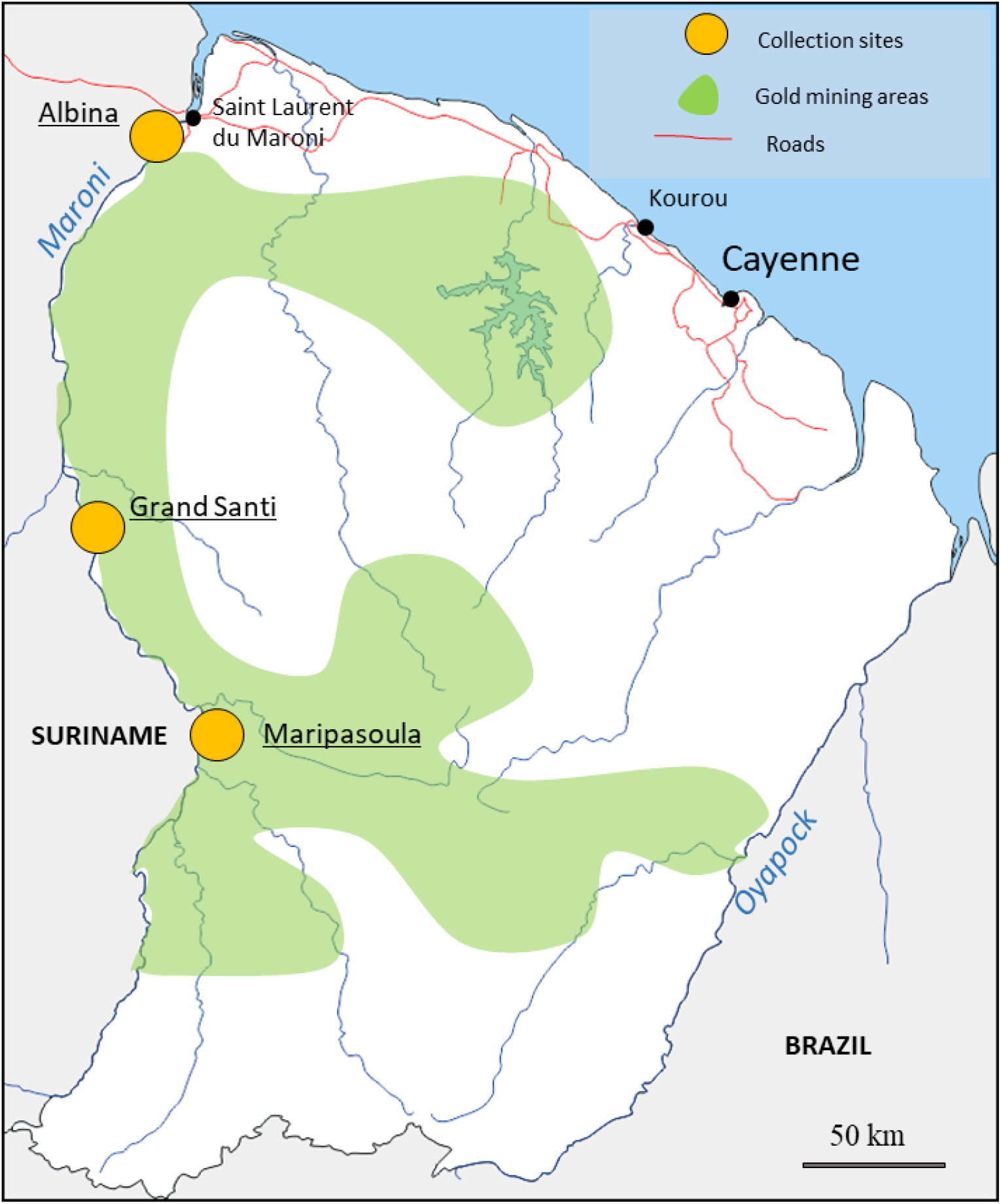
Sampling location of the Orpal study in French Guiana. Green area indicates illegal gold mining sites. Orange circles on the Maroni river indicate collection sites that were located in resting places.

**Figure 2.**
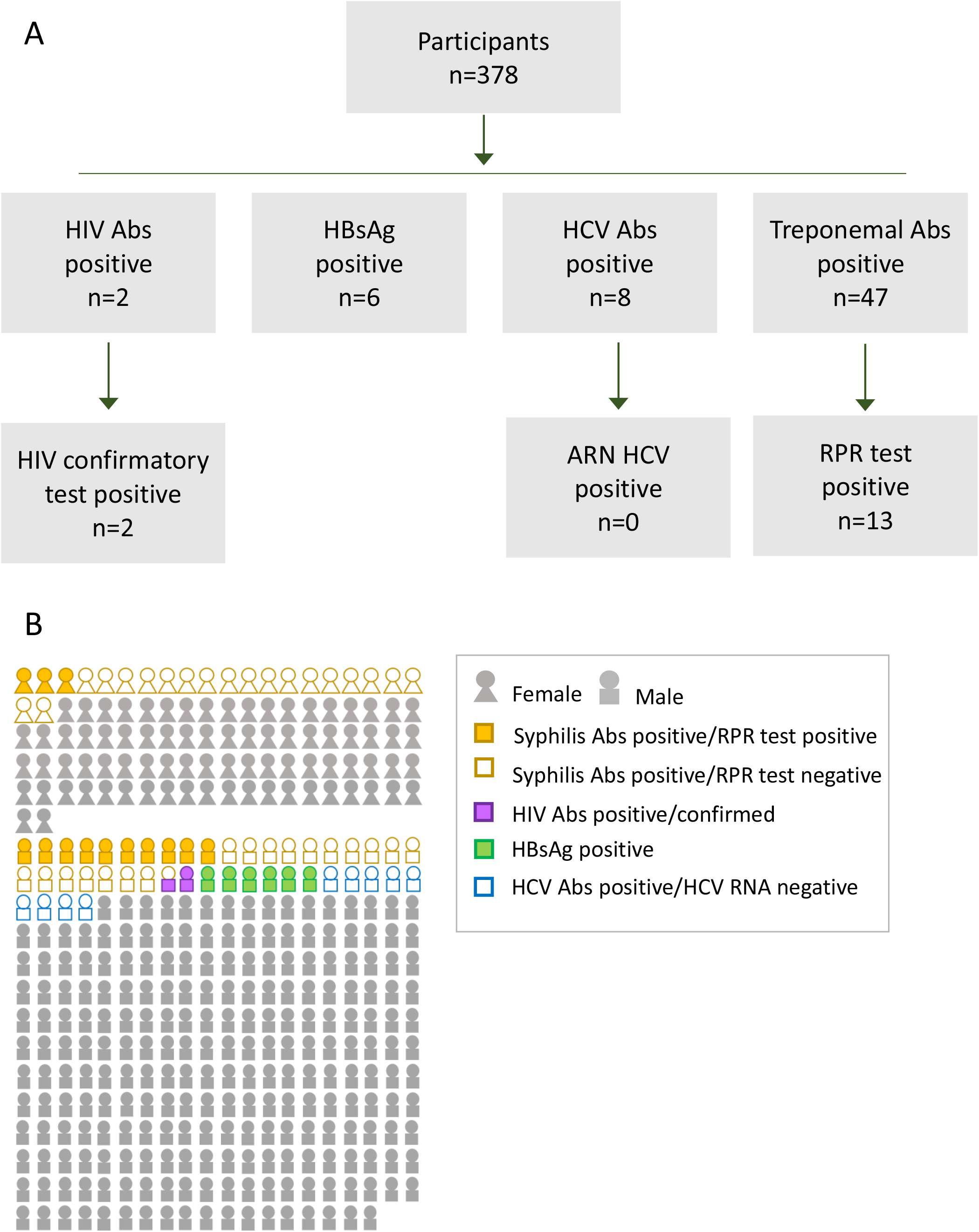
Screening of infections in illegal gold miners in French Guyana. A) Results for HIV, hepatitis B, hepatitis C, and syphilis in plasma samples HBsAg: hepatitis B surface antigen; Abs: antibodies; RPR: rapid plasma reagin test. B) Individual results.

**Figure 3.**
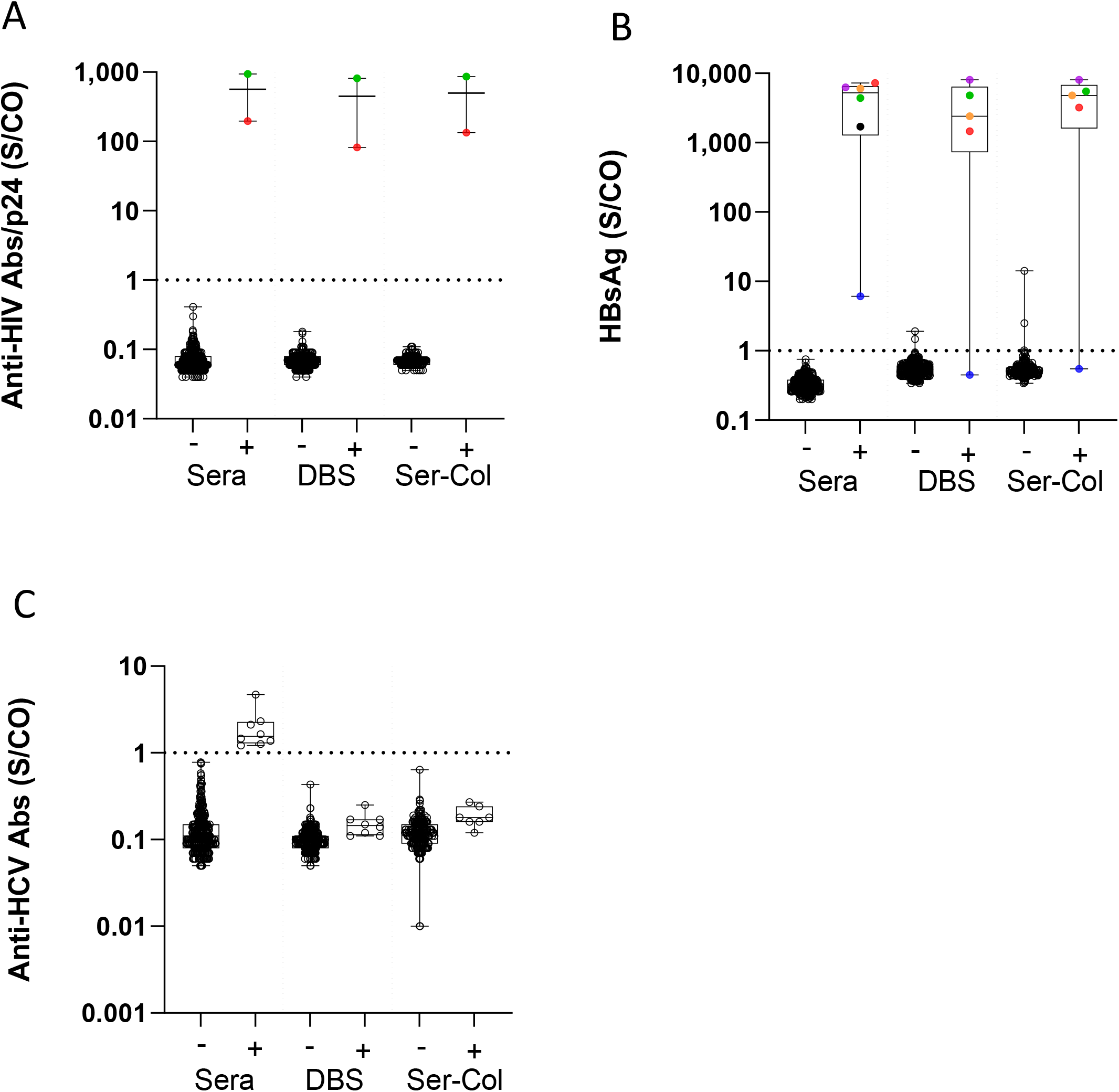
Results of HIV, hepatitis B and hepatitis C testing on Ser-Col and DBS samples. A) Results of HIV testing in Ser-Col samples and dried blood spot (DBS) according to HIV status in sera. B) Results of HBsAg testing in Ser-Col samples and dried blood spot (DBS) according to HBsAg status in sera. C) Results of hepatitis C antibody testing in Ser-Col samples and dried blood spot (DBS) according to anti-HCV status in sera. All samples tested positive for anti-HCV Abs were negative for HCV RNA.

**Figure 4.**
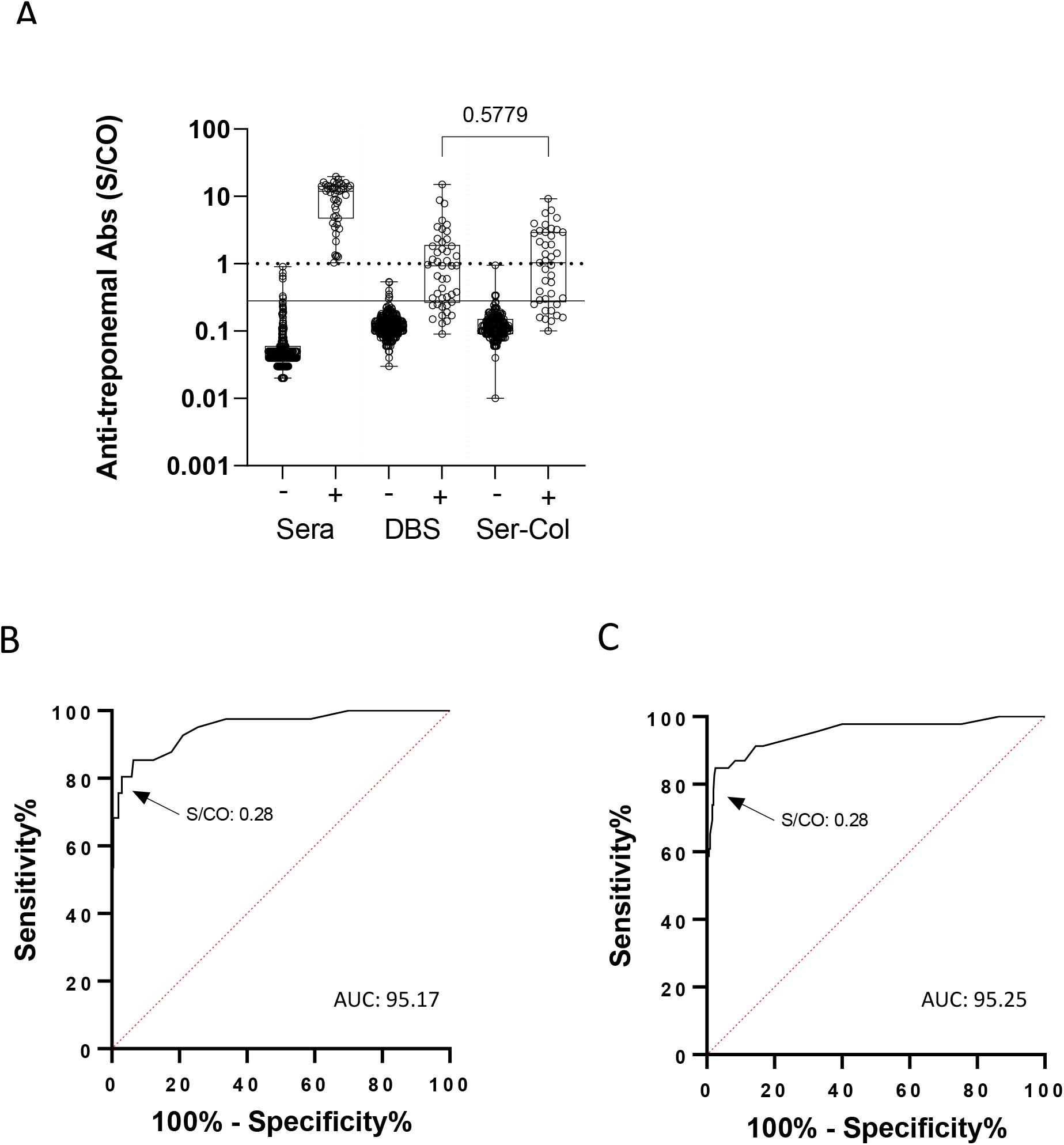
Anti-treponemal antibody results in Ser-Col and Dried Blood Spot samples. A) Anti-treponemal antibodies in Ser-Col samples and dried blood spot (DBS) according to syphilis serological status in plasma; dotted line: threshold of the manufactuer; solid line: optimized threshold for Ser-Col and DBS samples. B) Receiver operating characteristic curve (ROC) evaluating anti-treponemal antibody detection in Ser-Col samples. C) ROC evaluating anti-treponemal antibody detection in DBS samples.

### Sample processing

The processing of blood collected on the Ser-Col device (Labonovum, Netherlands) was performed using an automated system (SCAUT Ser-Col automation, Blok System Supply, Netherlands). The Ser-Col device contains a microfluidic paper separating plasma from whole blood. The automated system performed the elution from dried plasma contained in one centimeter of paper in 100µL of Ser-Col buffer.

Blood were spotted onto Whatman 903(tm) Protein Saver Cards (Whatman GmbH, Dassel, Germany) Elution were manual. One punch of 6 mm in diameter was eluted in 100µL of phosphate-buffered saline.

### Biological analysis

HIV p24/antibodies, HBsAg, anti-HCV antibodies, and anti-treponemal antibodies were assessed using fully automated chemiluminescence immunoassays (Alinity i, Abbott, IL, USA). Samples that tested positive for HIV were confirmed using a rapid confirmatory assay based on immune-chromatography and automated reading (Geenius(tm) HIV1/2 Confirmatory Assay, Bio-Rad, France). Plasma samples positive for anti-treponemal antibodies were tested by Rapid Plasma Reagin Assays (ASI RPR, Arlington Scientific Inc.). All assays were done in accordance with the manufacturer’s instructions.

### Data analysis and statistical methods

The HIV, HBV, HCV and syphilis status of participants was established by testing plasma samples. Ser-Col results were compared to plasma results, considered the gold standard for diagnostic testing in general, and to DBS results, considered the reference sampling method using dried blood. The Chi-square test was used to compare percentages. Receiver operating characteristic curves (ROCs) were used to identify the positive threshold of anti-treponemal antibodies in Ser-Col and DBS. Spearman’s non-parametric test was used to assess the correlation between results of anti-treponemal antibodies in Ser-Col and DBS, versus plasma. Data results were analyzed using GraphPad Prism 9.3.1 (GraphPad Prism Software Inc.).

## Results

We included 378 participants, 276 males and 102 females, in three illegal gold mining resting sites. The median (IQR) age of the participants was 39 years (31-48).

We first tested participants for HIV, HBV, HCV and syphilis infections using plasma to determine their infection status for these infections (Fig.1 and Table 1). Two participants tested positive for HIV. HIV infection was confirmed with the Geenius test. The pattern of antibodies showed IgG directed against gp160, gp41, p24 and p31 (low signal) of HIV-1 for the first participant, and gp160, gp41, and p24 with no p31 antibodies for the second one. We did not observe any reactivity for HIV-2 antigens (gp140, gp36). Six participants had HBV infection demonstrated by the presence of HBsAg. One participant had a weakly positive result with a signal-to-cut-off ratio (S/CO) of less than 10, corresponding to an HBsAg concentration of 0.5 IU/mL. Eight participants tested positive for anti-HCV antibodies, but all had index values close to the threshold of positivity. In addition, all of them tested negative for HCV RNA, suggesting a recovered hepatitis C infection or a false-positive result. 47 participants tested positive for syphilis using the treponemal test, (S/CO: 1.03 to 19.59). The rate of positivity for treponemal antibodies was higher in females than in males: 20/102 (19.6%) versus 27/276 (9.8%, p= 0.01), respectively. Among participants who tested positive for treponemal antibodies, 13 (27.6%) also tested positive using the non-treponemal test, suggesting an active stage of infection or a recently treated infection (3 females, 10 males). RPR dilution titers were 1:2 for five participants, 1:4 for six, 1:8 for one, and 1:32 for one.

**Table 1.**
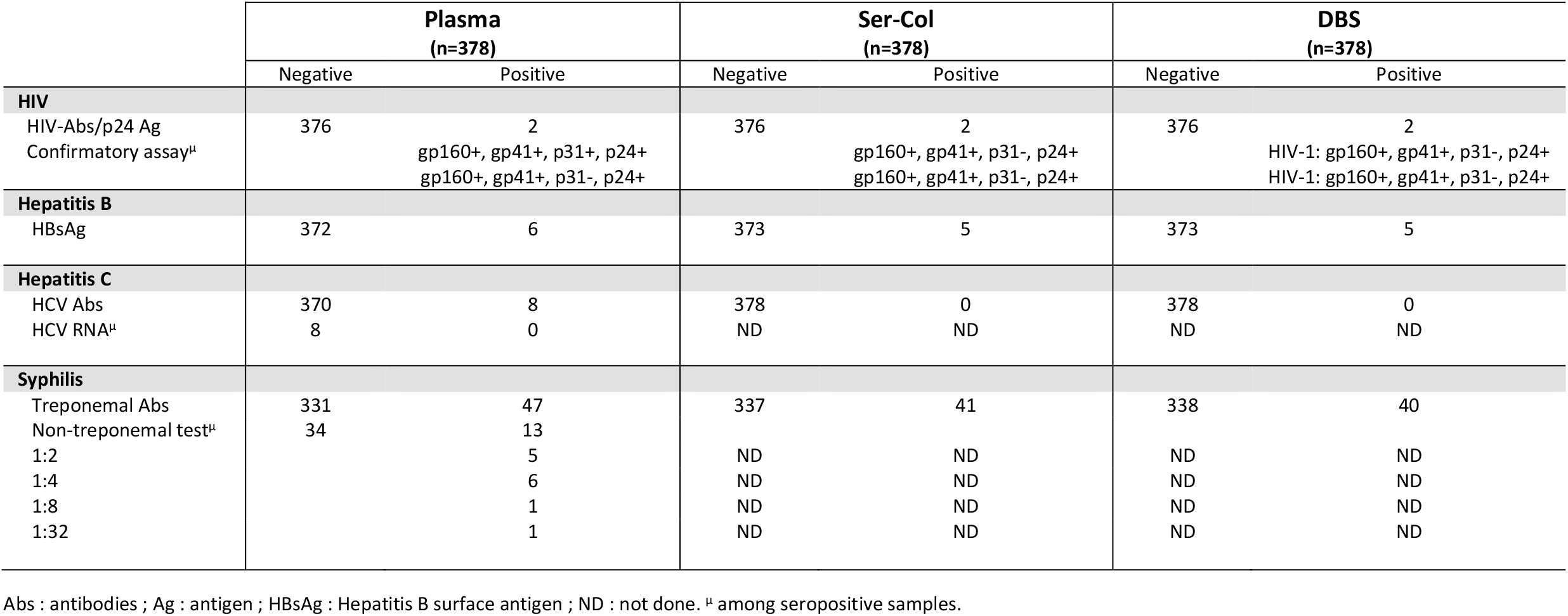
Serological results using plasma, Ser-Col, and DBS samples.

|  | **Plasma**<br>**(n=378)** |  | **Ser-Col**<br>**(n=378)** |  | **DBS**<br>**(n=378)** |  |
| --- | --- | --- | --- | --- | --- | --- |
|  | Negative | Positive | Negative | Positive | Negative | Positive |
| **HIV** |  |  |  |  |  |  |
| HIV-Abs/p24 Ag | 376 | 2 | 376 | 2 | 376 | 2 |
| Confirmatory assay<sup>u</sup> |  | gp160+, gp41+, p31+, p24+<br>gp160+, gp41+, p31-, p24+ |  | gp160+, gp41+, p31-, p24+<br>gp160+, gp41+, p31-, p24+ |  | HIV-1: gp160+, gp41+, p31-, p24+<br>HIV-1: gp160+, gp41+, p31-, p24+ |
| **Hepatitis B** |  |  |  |  |  |  |
| HBsAg | 372 | 6 | 373 | 5 | 373 | 5 |
| **Hepatitis C** |  |  |  |  |  |  |
| HCV Abs | 370 | 8 | 378 | 0 | 378 | 0 |
| HCV RNA<sup>u</sup> | 8 | 0 | ND | ND | ND | ND |
| **Syphilis** |  |  |  |  |  |  |
| Treponemal Abs | 331 | 47 | 337 | 41 | 338 | 40 |
| Non-treponemal test<sup>u</sup> | 34 | 13 |  |  |  |  |
| 1:2 |  | 5 | ND | ND | ND | ND |
| 1:4 |  | 6 | ND | ND | ND | ND |
| 1:8 |  | 1 | ND | ND | ND | ND |
| 1:32 |  | 1 | ND | ND | ND | ND |

Abs : antibodies ; Ag : antigen ; HBsAg : Hepatitis B surface antigen ; ND : not done. <sup>u</sup> among seropositive samples.

The Ser-Col samples then were tested for HIV, HBV, HCV and syphilis after automated elution. The eluates were assessed directly on the Abbott system without transfer into a secondary tube. All of the Ser-Col samples from HIV-seronegative participants tested negative, leading to a specificity of 100%. Two participants diagnosed with HIV infection were found positive using the Ser-Col device. The HIV confirmatory test was positive in the two samples. We observed a partial agreement between Ser-Col and plasma in one sample in whom the p31 band was detected in plasma but not in Ser-Col eluate (Table 1). Seven of the eight HBsAg carriers tested positive when using the Ser-Col device. The Ser-Col sample with the lowest HBsAg concentration in the paired plasma sample (0.5 IU/mL) tested negative. Two Ser-Col samples from HBsAg negative participants had signals above the cut-off and were considered false-positive. The S/CO were close to the positive threshold. All HCV seronegative participants were also found negative for anti-HCV antibodies using the Ser-Col device. The eight participants with the HCV Abs +/RNA-pattern were found negative using the Ser-Col device. All Ser-Col samples from participants seronegative for anti-treponemal antibodies in plasma tested negative. 20 out of 47 (42.5%) participants who tested positive for anti-treponemal antibodies in plasma had S/CO results below the threshold of the manufacturer. The signal on the cut-off index was significantly higher in participants who tested positive for anti-treponemal antibodies in the plasma. We established an optimized positive threshold based on a ROC curve. The best compromise was reached with an S/CO threshold of 0.28 instead of 1, corresponding to a sensitivity and a specificity of 74.6% and 98.4%, respectively. The AUC was 95.17 with this new threshold.

Finally, DBS samples were tested for HIV, HBV, HCV and syphilis. We observed identical screening results for HIV using DBS compared to plasma samples. The p31 band bands were also missing on the DBS samples of the first and second HIV positive participants (Table 1). As for the Ser-Col samples, the sample with the lowest HBsAg concentration tested negative using DBS. Two DBS samples collected from HBsAg negative participants had an S/CO above the positive threshold. These samples were not the same as those found false-positive using the Ser-Col samples. 23 out of 47 (48.9%) participants who tested positive for anti-treponemal antibodies in plasma had S/CO results below the threshold of the manufacturer. The ROC curve for the screening of anti-syphilis antibodies led to an AUC of 95.25, with a sensitivity and a specificity of 73.9% and 98.1%, respectively, with a threshold of 0.28.

## Discussion

Our study shows the feasibility of serological screening for sexually transmitted infections in a vulnerable population living in a remote area using a standardized dried blood device and a fully automated processing system. The clinical performance of the assays using the Ser-Col device appeared as good as those using DBS while overcoming some of the limitations related to DBS processing. This approach allows combined screening for HIV, HCV, HBV and syphilis infections on blood collected outside health care facilities in gold mining resting sites.

French Guiana has the highest prevalence of HIV and HBV among French regions (Moana et al., 2016; Nacher et al., 2018). The HIV, HBV, HCV and syphilis prevalences observed in our study are consistent with those previously reported in this population (Douine et al., 2019). With the exception of syphilis, the number of positive cases was relatively low, which leads to imprecise estimates of prevalence. These figures are close to HIV, HBV and HCV prevalence estimations in French Guiana. The prevalence of HIV has been estimated at 1.5% (Nacher et al., 2018), HBV 2.96% and HCV infection 0.67% (Moana et al., 2016). In remote areas and among vulnerable populations, some people living with HIV and viral hepatitis are unaware of their infection status (Organization, 2016). It is of primary importance to increase the frequency of screening for HIV, HBV and HCV infections. Capillary blood sampling and dried blood transportation could facilitate large-scale health surveys. This type of population testing has been carried out using DBS, for example for viral hepatitis (Meda et al., 2018) and SARS-CoV-2 (Warszawski et al., 2022).

This field study was carried out on difficult terrain. The population of illegal gold miners in French Guiana consists of migrants living far from transport networks and health infrastructure in a harsh tropical environment. In this study, sampling was done close to the field and in vitro diagnoses were performed in a central laboratory certified ISO 15189 for HIV, HBV, HCV and syphilis testing using a high-throughput serological platform. The transportation of samples from the field to a central laboratory is facilitated when using dried blood because there is no risk of the biological fluid spilling and no risk of virus transmission. Serological and molecular tests on DBS have shown satisfactory analytical performance for HIV and viral hepatitis (Lange et al., 2017a; Lange et al., 2017b). DBS is recommended by WHO for the diagnosis of HIV and viral hepatitis B and C to improve access to in vitro diagnosis of populations with poor access to plasma tests (Organization, 2017; World Health et al., 2014). Nevertheless, DBS suffers from significant limitations that hamper its use, including the lack of standardization of the sample and the absence of automation for the pre-analytical steps: punching, transfer and elution. The manual processing of DBS is a source of error and limits its integration in medical laboratories. The Ser-Col device and its automated processing overcome these limitations.

In our study, the screening on Ser-Col samples presented a high specificity, identifying HIV and HBV infections with the exception of one weakly HBsAg-positive sample. Two Ser-Col samples were false-positive for HBsAg but with low S/CO values. The specificity for HCV also appeared high but the sensitivity could not be calculated since all participants tested seronegative for HCV RNA. The performance of the syphilis assay is more uncertain since about a quarter of the low plasma positive participants were not detected when using the Ser-Col device. These participants had relatively weak positive results in plasma, suggesting an old, cured syphilis. However, a very recent infection would also be possible. Performance for syphilis screening was similar using the Ser-Col device and DBS. Even if the performance of screening for anti-treponemal antibodies on Ser-Col or DBS is lower than that of plasma, this method of blood sampling could be useful for syphilis testing in the field. A trade-off between the analytical performances of the test and its accessibility in the field should be considered to evaluate the benefit of in vitro diagnosis methods in the real world, as emphasized in the WHO guidelines (Organization, 2017; World Health et al., 2014).

Our study has several limitations. First, we did not evaluate loss of follow-up before participants obtained the results. This is an important aspect of screening strategies. Compared to POC tests, laboratory analyses, whether carried out on plasma or dried blood samples, entail a longer delay before the result is returned, and therefore a greater risk of patient loss to follow-up before the test results are given to the patient (Bottero et al., 2015). Another limitation is the low number of positive participants for HIV, HBV and HCV infections even though a large number of participants were tested. Finally, we only performed qualitative tests, whereas quantitative results may be interesting for the diagnosis of infectious diseases.

**In conclusion**, the Ser-Col device combined with automated elution allows the standardization and safety of the pre-analytical steps of analysis performed on dried blood samples. The clinical performances of the assays using the automated Ser-Col method appeared as good as those obtained with the DBS samples processed manually. Automated approaches to test capillary blood transported on dried blood devices are needed to enable testing of vulnerable populations living outside healthcare facilities and facilitate large-scale surveys assessing the burden of different infectious pathogens.

## Data Availability

Data are available on request to the corresponding author.

## Declarations

### Ethics approval and consent to participate

The study obtained approval from ethical committee in France: INSERM Ethics Evaluation Committee (No. 14-187 of 09.12.2014), in Suriname: CMWO. Approval No: VG 25-17; ClinicalTrials.gov identifier (NCT number: NCT03695770).

## Acknowledgements

The authors would like to thank all participants for participating in this study.

## CRediT authorship contribution statement

**Amandine PISONI** : Conceptualization, Methodology, Validation, Formal analysis, Writing - Original Draft ; **Elisa REYNAUD** : Validation ; **Maylis DOUINE** : Investigation ; **Louise HUREAU** : Investigation ; **Carmen ALCOCER CORDELLAT** : Investigation ; **Roxane SCHAUB** : Investigation ; **Dennis POLAND** : Conceptualization and Methodology ; Funding acquisition ; **Richard MONKEL** : Investigation : Resources ; **Joan LOMMEN** : Investigation ; **Konstantin YENKOYAN** : Conceptualization and Methodology ; Funding acquisition ; **Stephen VREDEN** : Writing - Review & Editing ; **Mathieu NACHER**: Investigation, Resources ; **Edouard TUAILLON** : Conceptualization and Methodology; Formal analysis: Resources ; Writing - Original Draft; Funding acquisition ; **All authors** : Writing - Review & Editing

